# Immune Modulation by Personalized vs. Standard Prehabilitation Before Major Surgery: A Randomized Controlled Trial

**DOI:** 10.1101/2025.06.03.25328894

**Authors:** Amélie Cambriel, Amy Tsai, Benjamin Choisy, Maximilian Sabayev, Julien Hedou, Elizabeth Shelton, Kreeti Singh, Jonas Amar, Valentin Badea, Serena Bruckman, Ed Ganio, Jakob Einhaus, Dorien Feyaerts, Ina Stelzer, Masaki Sato, Olivier Langeron, T. Adam Bonham, Dyani Gaudillière, Andrew Shelton, Cindy Kin, Brice Gaudillière, Franck Verdonk

**Author notes:** ***Corresponding author:*** Brice Gaudillière, MD, PhD. These authors contributed equally to this work (Amélie Cambriel, Amy Tsai, Benjamin Choisy, Maximilian Sabayev, Cindy Kin, Brice Gaudillière, Franck Verdonk).

## Abstract

**Importance:** Prehabilitation (prehab) programs are increasingly recognized for their potential to improve surgical outcomes. However, their efficacy remains debated, largely due to a lack of pathophysiologically-driven implementation and limited personalization.

**Objective:** To determine the impact of personalized versus standard prehab on preoperative physical, cognitive, and immune function and postoperative outcomes.

**Design, Setting, and Participants:** In this prospective, single-blinded, interventional trial conducted from October 2020 to April 2024 in a single academic medical center, 58 patients undergoing major elective surgery were randomized to standard (n=30) or personalized prehab (n=28) using block randomization.

**Intervention:** The personalized group received weekly remote coaching tailored to individual progress in four domains (nutrition, physical activity, cognitive training, and mindfulness), while the standard group followed a paper-based program without individualized support.

**Main Outcomes and Measures:** Primary clinical outcomes included cognitive assessments and physical performance measures, including the Wall Squat Test, Timed-up-and-go Test, 6-Minute Walk Test (6MWT). The primary immunological outcomes included major innate and adaptive immune cell frequencies and intracellular signaling responses measured using a 47-plex mass cytometry immunoassay.

**Results:** Fifty-four of 58 patients completed the study (n=27 per group). The personalized group exhibited significant improvements in physical measures (e.g., 6MWT; p=0.03) and fewer severe postoperative complications (4 vs. 11 Clavien-Dindo grade >1; p=0.04). Multivariable modeling identified profound and cell-type specific immune alterations post-prehab compared to baseline (AUROC=0.88 [0.79, 0.97], p=2-06; leave-one-out cross-validation), including dampened pERK1/2 signaling in classical monocytes and myeloid-derived suppressor cells after interleukin (IL)-2,4,6 stimulation, and reduced pCREB signaling in Th1 cells. In contrast, the standard group showed only moderate clinical improvements and no immune changes (AUROC=0.63, p=0.11).

**Conclusions and Relevance:** Our study demonstrates personalized prehab significantly altered the immunome before surgery, dampening inflammatory signaling responses previously implicated in the pathophysiology of key surgical outcomes, including surgical site infections and postoperative neurocognitive decline. These changes were accompanied by improved physical and cognitive function before surgery and decreased postoperative complications. Our findings support utilization of personalized prehab and provide an avenue for biologically-driven risk-stratification for patient selection, and individual tailoring of programs to optimize surgical readiness and recovery.

**Trial Registration:** NCT04498208

**KEY POINTS:** 

**Question:** How do personalized prehabilitation programs modulate the peripheral immune system in patients undergoing major elective surgery?

**Findings:** In a randomized trial, patients scheduled for major elective surgery received either personalized or standard prehabilitation. High-dimensional immune profiling with mass cytometry revealed profound and cell type-specific dampening of pro-inflammatory signaling responses in the personalized prehabilitation group (AUROC=0.88, n=27), but not in the standard group (AUROC=0.63, n=27). Patients in the personalized prehabilitation group also showed significant improvements in both physical and cognitive function, with significantly fewer severe postoperative complications (4 vs. 11).

**Meaning:** Personalized prehabilitation dampens patients’ pre-operative inflammatory state and enhances recovery by improving physical and cognitive outcomes, suggesting tailored interventions may optimize surgical preparedness and reduce complications.

## INTRODUCTION

Up to 30% of patients undergoing major elective surgery suffer postoperative complications,^1,2^ leading to delayed functional recovery, prolonged hospital stays,^3,4^ and significant healthcare costs,^4^ exceeding $145 billion annually in the United States.^5^

In recent decades, the adoption of enhanced recovery after surgery (ERAS) protocols has significantly improved surgical outcomes via interventions from the day before surgery to hospital discharge.^6^ By contrast, prehabilitation (prehab) focuses on enhancing patients’ physiological reserve weeks to months before surgery in order to improve their resilience to surgical trauma.^7^ While early prehab programs were focused on physical exercise,^8^ more recent multi-modal approaches, including some combination of exercise, nutrition, cognitive training, and stress reduction, have shown more promise in improving surgical recovery.^7,9–11^ Amidst the growing body of literature on prehab, the true impact on postoperative outcomes remains debated. Recent meta-analyses of studies conducted between 2000 and 2018 report conflicting results, particularly regarding postoperative complications and length of stay.^10,12–17^ One key reason for this variability is the heterogeneity of prehab programs, the degree of personalization and coaching, and patient selection criteria.^10,12^ In a healthcare system bound by increasing resource constraints, there is a critical need to identify patients most likely to benefit from prehab and to adapt programs to their needs.

Current tools for risk-stratification, such as the Risk Analysis Index-Administrative (RAI-A), the American College of Surgeons National Surgical Quality Improvement Program (ACS NSQIP) surgical risk score, and Fried’s frailty score, rely exclusively on clinical data,^18–21^ and have limited predictive performance. A better understanding of the pathophysiological effect of prehab programs on mechanisms driving surgical recovery is a critical first step to design effective prehab programs tailored to each patient’s biology.

Surgical trauma triggers profound local and peripheral immune responses engaging a network of innate and adaptive immune cells.^22–27^ Successful surgical recovery requires a functional balance between pro-inflammatory and immunosuppressive responses essential for pathogen defense, wound healing, and pain resolution.^28–30^ As such, in-depth immune profiling before and after prehab holds promise for identifying modifiable immunologic parameters to inform the design of personalized prehab interventions.

Here, we employed high-dimensional mass cytometry to comprehensively and dynamically characterize the effect of a personalized prehab regimen on 1,096 innate and adaptive immune cell frequency and signaling features (the immunome) measured in patient blood samples collected before surgery. We applied Stabl^31^, a sparse machine learning algorithm combining multivariable modeling and data-driven feature selection, to identify immune features distinguishing patients before and after prehab. The analysis identified a profound immune-modulating effect of personalized prehab pinpointing critical cell-type and signaling-specific immune alterations that have been previously implicated in surgical recovery.^31–33^

## MATERIALS AND METHODS

### Trial design and ethics statements

This prospective, single-center, single-blinded, randomized controlled trial compared standard versus personalized prehab in patients undergoing major elective surgery. It was conducted in accordance with the declaration of Helsinki, approved by Stanford’s Institutional Review Board (IRB-57570), and registered in 2020 on ClinicalTrials.gov (NCT04498208). The study follows the CONSORT guidelines.

### Participants

Eligible participants were ≥18 years old, scheduled for major elective surgery (abdominal, thoracic, plastic and reconstructive, and neurosurgical procedures) under general anesthesia at Stanford Health Care, with surgery planned ≥14 days after enrollment. Inclusion required fluency in English and ability to provide informed consent. Exclusion criteria included premorbid conditions or orthopedic impairments contraindicating exercise, cognitive disabilities defined as evolutive neurological or neurodegenerative disease, expected length of stay in the hospital <48h, American Society of Anesthesiologist (ASA) score ≥4, terminal illness under palliative care, illiteracy, legal conservatorship, pregnancy or breastfeeding, and missing pre- or post-prehab blood samples. The study design is represented in **Figure 1A**.

**Figure 1:**
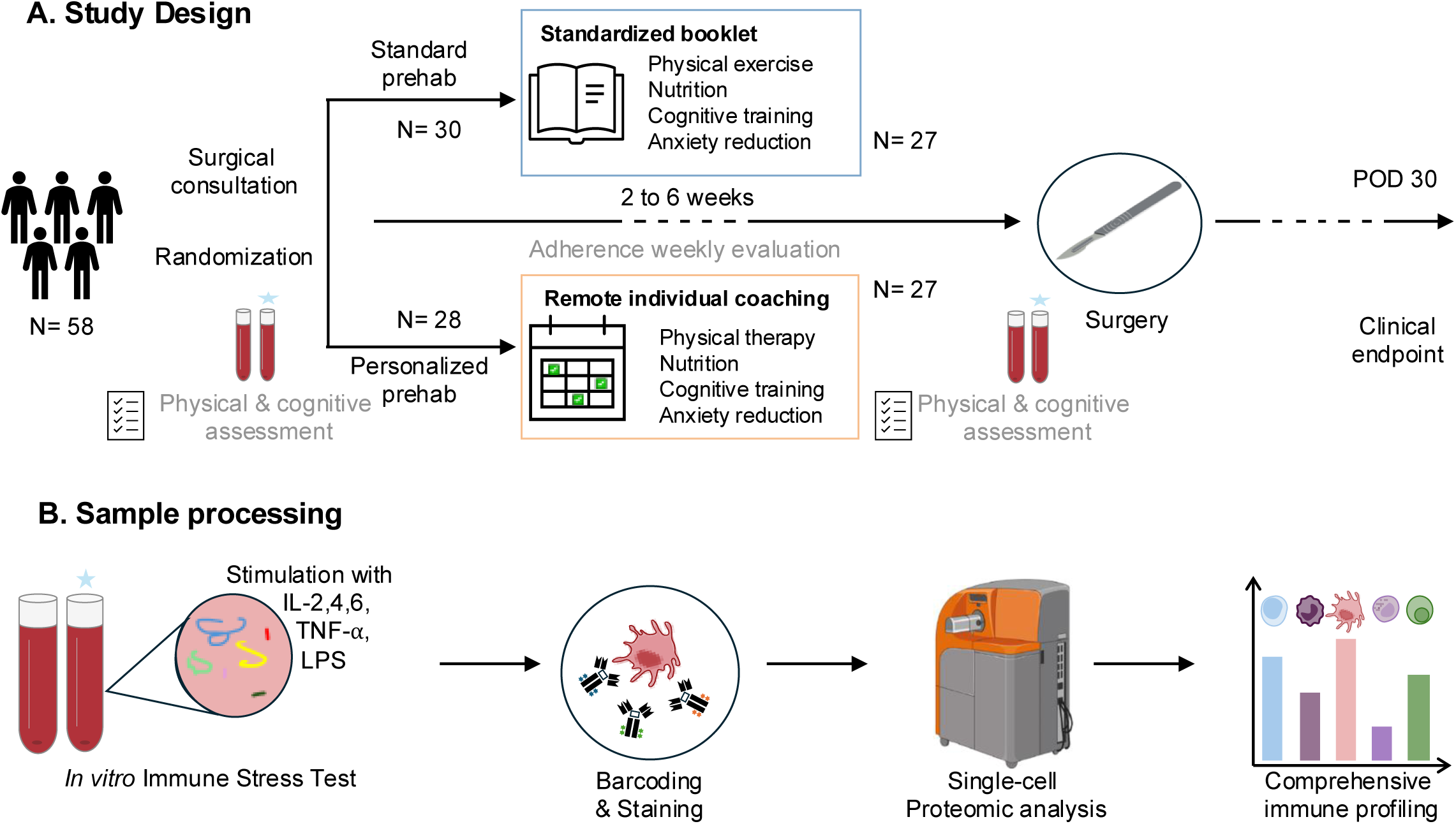
Study design and workflow. **A.** Study design: 58 patients undergoing major surgery were randomized to receive either standard prehabilitation (prehab) (n=30) or personalized prehab (n=28) at the preoperative surgical consultation. Clinical evaluation and blood sample collection was performed at two time points: at time of randomization (pre-prehab) and preoperatively (post-prehab). Prehab lasted 2-6 weeks, and adherence was evaluated weekly. 27 patients from each arm completed prehab and were included in the final analysis. Clinical endpoints included complications assessed within 30 postoperative days (POD) using the Clavien-Dindo classification and hospital length of stay. **B.** Sample processing: whole blood samples were either left unstimulated to measure endogenous intracellular activities or exogenously stimulated with toll-like receptor-4 agonist lipopolysaccharide (LPS), Interleukins (IL)-2,4,6, or tumor necrosis factor-alpha (TNF-⍺) to activate canonical pathways in a surgical trauma model and provide an in-depth analysis of patients’ immunomes.^33,53^ Following erythrocyte lysis, samples were barcoded and stained with both surface and intracellular antibodies according to standard protocols and assessed simultaneously to minimize experimental variability.^26,27^ Samples were measured using a 47-multiplex mass cytometry assay yielding a highly comprehensive immune profile for each patient.

### Randomization and blinding

Patients were randomized in blocks of 10, stratified by age, to receive standard or personalized prehab two to six weeks before surgery. Outcomes assessors, surgeons, anesthesiologists, and statisticians were blinded to group allocation. Patients were not blinded to prehab assignment due to the nature of the intervention.

### Prehab programs

In the standard prehab group, patients received a printed booklet with guidance on physical exercise, nutrition, stress-reduction, and cognitive training. Physical exercise recommendations included four difficulty levels of strength and cardio exercises and seven stretching exercises. Strength exercises included upper body, lower body, and core. Cardio exercise regimens included low-intensity and high-intensity options. The nutritional recommendations followed a Mediterranean-style diet, stress and pain management included *the Breathe-Scan-Visualize- Check-in* technique, and cognitive training recommendations included daily use of the Lumosity® app. After one orientation session with a physician member of the research team reviewing the booklet, patients were instructed to select their highest exercise level based on their comfort and received no further coaching.

Patients in the personalized prehab group attended twice-weekly one-on-one remote coaching sessions tailored to their individual abilities and progress, including the same four domains as the standard prehab. Patients had access to an online platform (Notion®) that offered daily meditation recordings, explanations for the exercise program of the week, daily recommendations for healthy meals and Mediterranean-diet recipes, a reminder to practice cognitive training on the Lumosity® app, as well as a feedback section where patients could record their challenges to discuss during coaching sessions. Sessions were provided weekly by both a physician, who focused on nutrition, cognition, and behavioral strategies, and a physical therapist, who specialized in exercise. Initial sessions were dedicated to education and creation of a tailored prehab program. Subsequent sessions were focused on patients’ achievements and resultant adaptions of the prehab plan. If surgery was scheduled more than six weeks after randomization, patients were asked to continue the program, but no remote coaching sessions were scheduled after six weeks.

### Data Collection

Whole blood collection, for immune single-cell analysis, and physical and cognitive ability assessments occurred at two time points: baseline (pre-prehab) and preoperatively (post-prehab). A trained member of the research team, blinded to group assignment, administered the 6-Minute Walk Test (6MWT), the Timed up-and-go Test, Wall Squat Test, and Quick Mild Cognitive Impairment (qMCI) Test. Adherence was monitored weekly by a blinded evaluator using a 4-point Likert scale (1 = no prehab engagement, 4 = daily adherence). Study data were collected and managed using REDCap electronic data capture tools hosted at Stanford University.^34^

### Study outcomes

Primary outcomes included the effect of standard versus personalized prehab on 1) improvement in physical and cognitive performance before surgery, 2) 30-day postoperative complications (using the Clavien-Dindo classification^35^) and hospital length of stay, and 3) immunological measures.

### Single-cell analysis

Collection, processing, and analysis of blood samples by mass cytometry followed previously described protocols^27,32^ (Supplementary Methods).

### Statistical analysis

Statistical analyses were performed using Python3® and the following libraries: NumPy (v1.26.2), pandas (v2.1.4), matplotlib (v3.8.2), seaborn (v0.13.0), SciPy (v1.11.4), and scikit-learn (v1.3.2). Analysis code and datasets are available upon request.

#### Power analysis

Power calculations indicated a sample size of 29 patients per group was required, based on an estimated area under the receiver operating characteristic curve (AUROC) of 0.85, a proportion of patients in each group of 0.50, a confidence interval width of 0.25, a 90% confidence level, and an expected 5% loss to follow-up.

#### Analysis of clinical outcomes

We assessed within-group changes in physical, cognitive, and immune function following prehab. Comparisons of pre- versus post-prehab scores in each group were performed using a Wilcoxon signed-rank test for the qMCI, 6MWT, Timed-up-and-go Test, and Wall Squat Test.

Weekly adherence scores were averaged over the prehab period and compared between groups using the Mann-Whitney U test. Postoperative complications within 30 days (Clavien-Dindo classification) and hospital length of stay between groups were also compared with a Mann-Whitney U test. If patients presented more than one complication, the most severe complication grade was used to compare grade severity between groups.

#### Immune profile Classification

Following preprocessing and visualization of the single-cell proteomic dataset (**Supplementary Methods**), we applied a multivariable modeling approach to identify differences in patients’ immune profiles encompassing endogenous intracellular signaling activities, signaling responses to extracellular stimulations, and immune cell frequency before and after the prehab regimen for each group. To identify a set of distinguishing immune features between time points (pre- and post-prehab), the Stabl algorithm (**Supplementary Methods**),^31^ was employed to fit a classification model. Predictive performance was evaluated using the AUROC, computed through a leave-one-out cross-validation approach tailored for multi-omic data integration.

## RESULTS

### Study population

Fifty-eight patients undergoing major elective surgery were randomized to receive either standard (n=30) or personalized (n=28) prehab 2-6 weeks before surgery, between June 2020 and September 2022 (**Figure 1**, CONSORT diagram **Figure S2**). Patients in the standard group followed a booklet-based exercise, nutrition, stress reduction, and cognitive training regimen. Patients in the personalized prehab group received tailored recommendations through twice-weekly remote coaching sessions.

Three patients in the standard group and one in the personalized group withdrew, resulting in 27 patients per group with complete paired blood samples for analysis (**Figure S2**). Baseline characteristics (**Table 1**) were comparable between groups in terms of age, gender, and ASA score. Most patients (79.6%) underwent abdominal surgery (n=19 in the standard group and n=24 in the personalized group, p=0.05), and 50.0% had cancer in both groups.

**Table 1:** Demographic characteristics of the study population.

|  | All<br>n=54 | Standard prehab<br>n=27 | Personalized prehab<br>n=27 | p |
| --- | --- | --- | --- | --- |
| **Age, years** | 57.00 [45.00,66.75] | 52.00 [45.00,69.00] | 60.00[45.00,66.00] | 0.68 |
| **Gender, male** | 23 (43.0) | 12 (44.44) | 11 (40.74) | 0.42 |
| **BMI (kg/m<sup>2</sup>)** | 25.07 [22.25,30.40] | 25.11[22.29,30.11] | 24.51[22.58,30.83] | 0.93 |
| **ASA, class** |  |  |  | 0.07 |
| I | 2 (3.70) | 1 (3.70) | 1 (3.70) |  |
| II | 23 (43.0) | 8 (29.62) | 15 (55.56) |  |
| III | 28(51.9) | 17 (62.96) | 11 (40.74) |  |
| IV | 0 (0) | 0(0) | 0(0) |  |
| **Surgery type** |  |  |  | 0.05 |
| **Abdominal surgery** | 43 (79.6) | 19(70.37) | 24 (88.89) |  |
| Obesity surgery (sleeve or bypass) | 8 | 4 | 4 |  |
| Colectomy | 15 | 4 | 11 |  |
| Abdominal debulking | 13 | 8 | 5 |  |
| Pancreatic surgery | 2 | 1 | 1 |  |
| Esophagectomy | 2 | 1 | 1 |  |
| Ileostomy take down/appendectomy | 4 | 2 | 2 |  |
| **Urologic procedure** | 3 (5.56) | 3 (11.11) | 0 |  |
| Prostatectomy | 1 | 1 | 0 |  |
| Cystectomy | 1 | 1 | 0 |  |
| Renal transplant | 1 | 1 | 0 |  |
| **Thoracic (lobectomy)** | 2 (3.70) | 2 (7.41) | 0 |  |
| **Major maxillofacial surgery** | 6 (11) | 3 (11.1) | 3 (11.1) |  |
| **Charlson comorbidity score** | 4[2,5] | 4[1.75,5] | 2[2-5] | 0.75 |
| **Comorbidity** |  |  |  |  |
| **Myocardial infarction or ischemic arteriopathy** | 10(18.52) | 6 (21) | 4(14.81) | 0.09 |
| **Congestive heart failure** | 1 (1.85) | 0 | 1 (3.70) | 1 |
| **Chronic pulmonary disease** | 9(16.67) | 7(25.93) | 2(7.41) | 0.05 |
| **Diabetes** | 7(12.96) | 2(7.41) | 5(18.52) | 0.23 |
| **Renal disease\*,** | 5(9.25) | 2(7.41) | 3(11.11) | 1 |
| **Cancer, localized** | 27(50.00) | 16(59.26) | 11(40.74) | 0.28 |
| **Cancer, metastatic** | 2(3.70) | 0 | 2(7.41) |  |
| **Prehab duration, days** | 32.5[23.5,42.0] | 35.0[22.0,42.0] | 31.0 [28.0,42.0] | 0.89 |

Values are expressed as medians [IQR] or numbers (%)
\*Renal disease is defined as creatinine >3 mg/dL

The median prehab duration was 35 [22,42] days in the standard group and 31 [28,42] days in the personalized group (p=0.89). Adherence on a 4-point Likert scale was higher in the personalized group (mean 3.3 ± 0.4) compared to the standard group (mean 3.0 ± 0.54; p = 0.03).

#### Personalized prehab improves preoperative cognitive and physical function and reduces postoperative complication severity

Functional evaluation after prehab revealed that patients in the personalized prehab group significantly improved both their physical function (6MWT in meters: 496 [340 ,619] pre-prehab vs. 546 [350,728] post-prehab, p=0.05; timed up-and-go test in seconds: 9.0 [6.7,9.9] pre-prehab vs. 6.9 [5.3,8.5] post-prehab, p=0.01; and wall squat time in seconds: 37.4 [26.3,59.2] pre-prehab vs. 66.2 [34.2,92.8] post-prehab, p<0.001) as well as cognitive function (qMCI score 66 [56 ;74] pre-prehab vs. 72 [65,76] post-prehab, p=0.03). In contrast, patients from the standard prehab group only improved in the 6MWT (507 [368; 623] pre-prehab vs. 573 [425, 707] post-prehab p=0.03), with no significant gains in other metrics (**Figure 2**).

**Figure 2:**
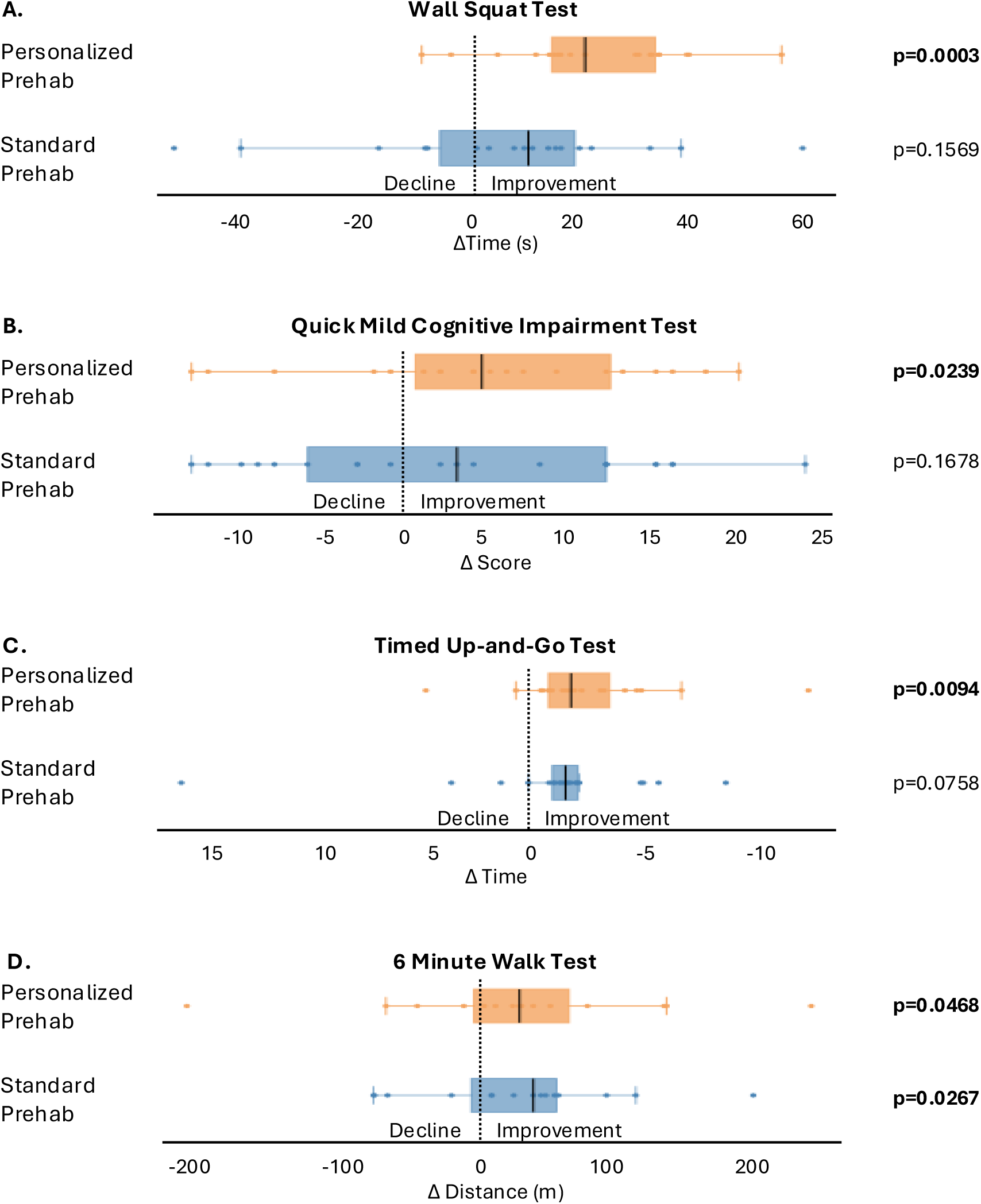
Physical and cognitive assessment evolution after prehab. Barplots depicting the median and IQR value of the score difference between pre- and post-prehab in each group for the **A.** Wall Squat Test, **B.** Quick Mild Cognitive Impairment Test, **C.** Timed-Up-and-Go Test, and **D.** 6 Minute Walk Test. Individual values are represented as dots.

Significantly fewer patients in the personalized group experienced moderate to severe postoperative complications (Clavien-Dindo classification grade >1^35^) compared to the standard prehab group (n=4 vs. 11 respectively, p=0.04, Mann Whitney U test, **Table S2)**. No difference was observed in hospital length of stay (2.1 [0.7, 4.0] vs 2.6 [1.5, 6.0], respectively; p=0.48).

#### Single-cell intracellular signaling activities differentiate patients before and after prehab in the personalized prehab group

To investigate whether prehabilitation modulates patients’ immunome, a 47*-*parameter mass cytometry assay was employed to extract single-cell immune features from blood samples collected before and after prehab. Uniform Manifold Approximation and Projection (UMAP) analysis was performed to provide a visual synopsis of all single-cell immune features measured, including major innate and adaptive immune cell subset frequencies, their endogenous intracellular activities (e.g., phosphorylation states), and capacities of each population to respond to a series of extracellular immune perturbations, including lipopolysaccharide (LPS), tumor necrosis factor (TNF)-α, and a combination of interleukins (IL)-2,4,6 (**Figure 3**). This approach provided a single-cell atlas of immune cell type-specific changes in intracellular signaling states in response to prehab (**Figure 3**).

**Figure 3:**
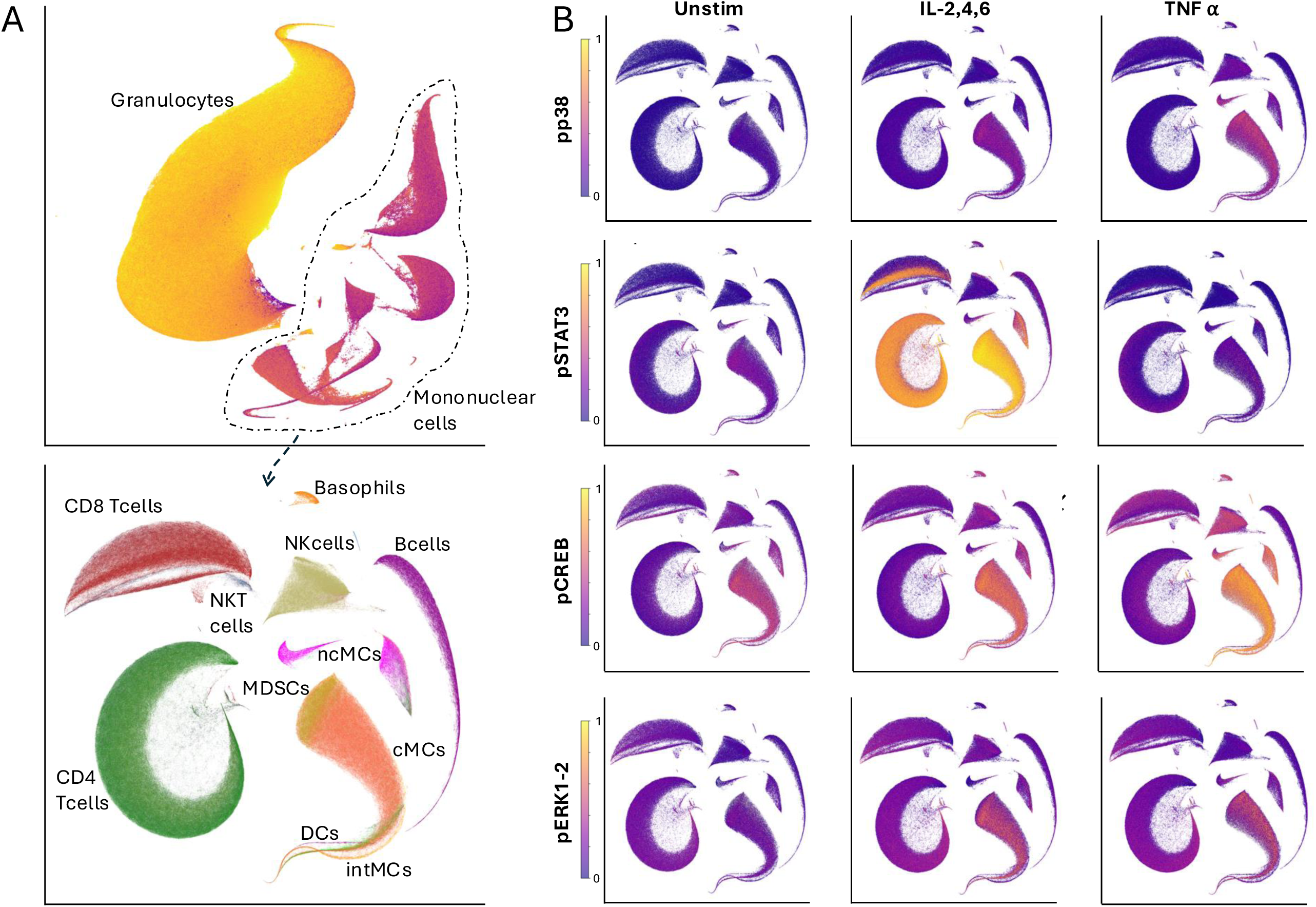
In vitro stress test before surgery analyzed with mass cytometry reveals cell-type- and signaling-specific responses. **A.** Uniform manifold approximation and projection (UMAP) representation of the single-cell mass cytometry dataset. *Upper panel*: all live leukocytes, including neutrophils and mononuclear cells; *lower panel*: UMAP representation of mononuclear cells only. UMAPS are clustered by cell type and annotated. **B.** UMAPs representing all mononuclear cells colored according to levels of intra- cellular signaling in endogenous state (‘Unstim’) and after in vitro stimulation with IL-2,4,6 and TNF-⍺. *Abbreviations: cMCs: classical monocytes; CREB: cAMP-response element binding protein; IL: Interleukin; intMCs: intermediate monocytes; mDCs: myeloid dendritic cells; MDSCs: myeloid-derived suppressor cells; ncMCs: non-classical monocytes; NK: natural killer; NKT: natural killer T cells; pDCs: plasmacytoid dendritic cells; ERK: Protein Kinase R-like ER Kinase; p: phosphorylated; JAK/STAT: Janus kinase/signal transducer and activator of transcription; TNF-*⍺*: Tumor Necrosis Factor alpha*

Immune cell subsets were further evaluated using an established manual gating strategy, yielding a total of 1,096 immune cell frequency and signaling features. The sparse machine learning algorithm, Stabl,^31^ was employed to determine whether patients’ immunome differed after prehab compared to baseline in each arm. The Stabl analysis identified a robust multivariable model that accurately differentiated samples collected before and after prehab in the personalized prehab group (AUROC = 0.88 [0.79, 0.97]; p=2e-06). In contrast, the multivariable analysis did not successfully differentiate samples before and after prehab in the standard prehab group (AUROC= 0.63 [0.48, 0.78], p=0.12). Between-group comparison of immune difference before and after prehab did not yield a statistically significant model (AUROC 0.58 [0.41,0.73], p=0.34).

Analysis of features selected by the Stabl model revealed cellular elements of a patient’s immunome that were the most reliably altered by personalized prehab (**Figure 4**, **Figure S3**). In adaptive cell types, personalized prehab resulted in decreased endogenous pCREB signaling in Th1-naïve and MAIT CD8^+^ T cells, with increased reactivity to exogenous TNF-⍺ stimulation, as well as a decreased endogenous pSTAT6 signaling in CD4^+^ Terminally Differentiated Effector Memory T cells. In innate immune populations, personalized prehab decreased pERK signaling in response to IL-2,4,6 in myeloid-derived suppressor cells and classical monocytes, as well as pMAPKAPK2 signaling in response to TNF-⍺ in intermediate monocytes.

**Figure 4:**
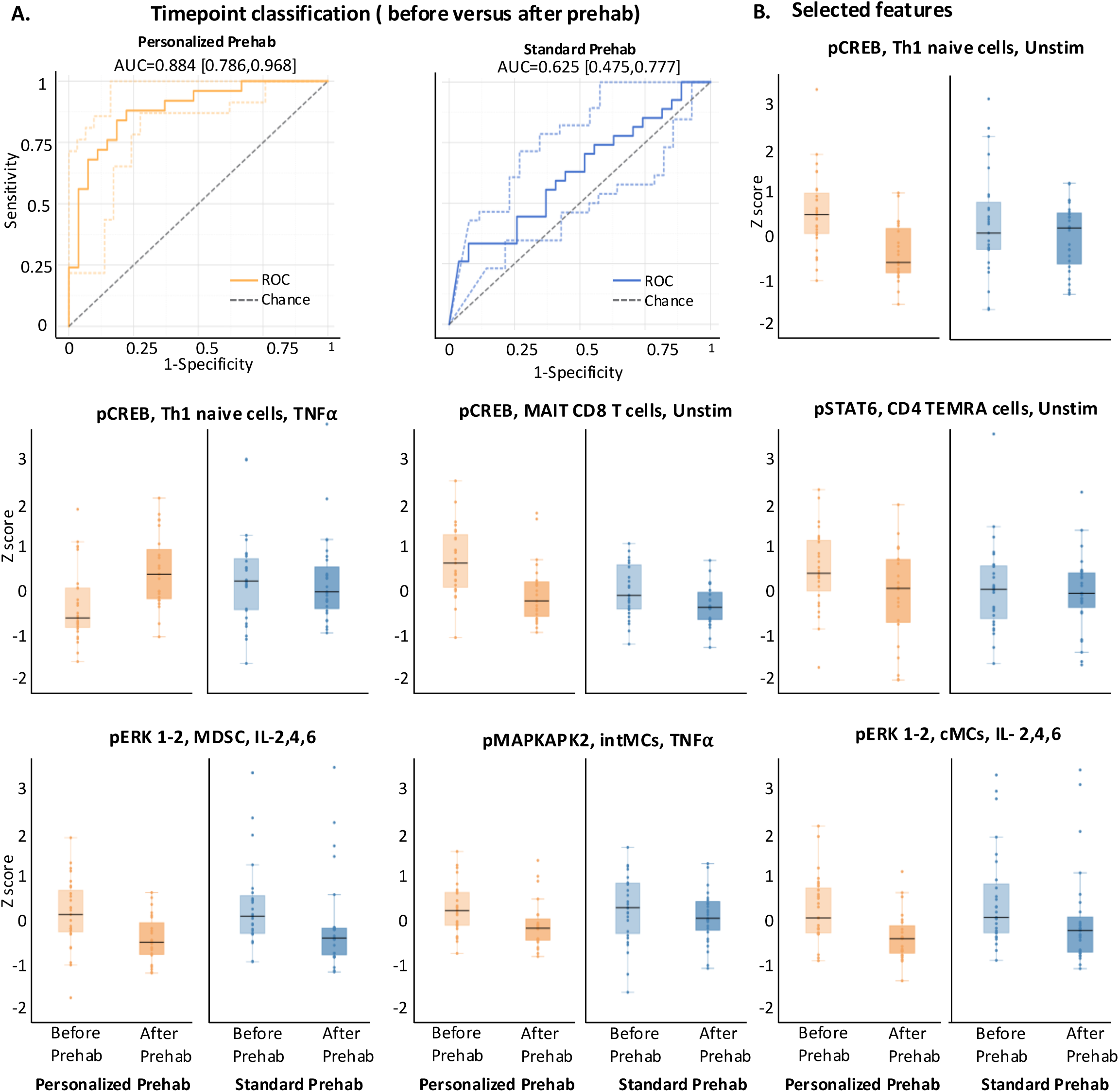
Multivariable classification of immune profiles before and after prehab. **A.** Classification performance of the Stabl multivariable model for identifying immune modifications after prehab in both groups. **B.** Boxplots depict the median and interquartile range of the single-cell mass cytometry features selected by the Stabl model. Abbreviations: *MAIT: Mucosal-Associated Invariant T cells; MAPKAPK2: Mitogen-activated protein kinase-activated protein kinase 2; TEMRA: Terminally Differentiated Effector Memory T cells*

These findings suggest that personalized, but not standard, prehab exerts a profound immunomodulatory effect on the immunome. Specifically, the results show that personalized prehab induces a dampening of basal JAK/STAT signaling in adaptive immunity and a decrease in MyD88 reactivity to stimulation in innate cells indicative of a reduction of the basal inflammatory state after prehab.

## DISCUSSION

In this randomized controlled trial, we conducted an extensive analysis of the effects of standard versus personalized prehab on patients’ immunological, physical, and cognitive function before major surgery. Our study shows that patients following a remotely delivered, personalized prehab regimen had increased adherence, improved physical and cognitive function after prehab, and less severe postoperative complications. Single-cell intracellular signaling analyses further revealed that personalized, but not standard, prehab significantly modulated patients’ immunomes, with a dampening of basal JAK/STAT and MyD88 signaling in adaptive immune cells, and a reduction of both basal and stimulated MyD88 pathway activity in innate immune cells, suggesting a reduction in pro-inflammatory pathways.

From a clinical standpoint, our results dovetail with previous studies examining the impact of prehab on postoperative outcomes, showing that prehab increases patients’ physiological reserve and improves recovery.^36^ Improvement in physical, cognitive, and postoperative outcomes were more pronounced in the personalized than the standard prehab group, supporting the efficacy of providing coaching support tailored to each patient during their preoperative journey. Notably, the personalized prehab program was delivered entirely through remote coaching of patients as our trial was conducted during the COVID-19 pandemic. As reported in other studies, patient satisfaction and adherence remained strong throughout the program.^37^ Previous studies have found that patients prefer remote administration over onsite prehabilitation^17^ as home-based programs minimize logistical barriers, reduce the burden on caretakers, and are more approachable, all of which likely contributed to our observed outcomes.^17,38^ Another key factor that contributed to the superior efficacy of personalized prehab over standard prehab is the higher adherence rate. The twice-weekly individual remote coaching with a physical therapist and a physician facilitated the development of a realistic plan for progress in each domain, as well as an element of personal accountability. This approach placed patients in a positive feedback loop, helping them recognize their accomplishments and fostering motivation.^39^

Recent meta-analyses on prehab efficacy highlight the necessity of targeting high-risk patients and monitoring intervention effectiveness before surgery.^10,40^ Our findings support this perspective: the more resource-intensive, coached version of prehab yielded significant physical, cognitive, and immunological improvements. These outcomes corroborate the need and provide an avenue for risk-stratification and progress monitoring.

Notably, several key immune features modulated by personalized prehab in our study resonate with features previously associated with surgical site infection^31,33^ and postoperative neurocognitive decline,^32^ suggesting they are interesting candidates for immune-based risk diagnostics and prehab monitoring. For example, patients at risk for surgical site infection display an exacerbated MyD88 response after surgery^31,33^ which is dampened by personalized prehab as we observe a reduction in MAPKAPK2 signaling in response to an exogenous stressor (TNF-⍺) in intermediate monocytes and pERK signaling in response to IL-2,4,6 in classical monocytes and myeloid-derived suppressor cells. Similarly, increased pCREB response in adaptive cells following personalized prehab may be clinically meaningful. Patients at risk for postoperative cognitive decline exhibit dampened pCREB response in adaptive cells to exogenous stress, this indicates that prehab may mitigate postoperative cognitive decline risk after surgery.^32^ Interestingly, dampened MyD88 response to exogenous stress, is a hallmark of immunosenescence,^41,42^ a dynamic and multifactorial process affecting both innate and adaptive immune cells. It also plays an important role in inflammaging and frailty seen in patients at risk for postoperative cognitive decline^32^ and other frailty-related postoperative complications.^43–46^ Our results show that prehab helps to mitigate this ill-adapted immune response, corroborating other works that have shown an improvement in immune function with preoperative exercise and nutrition optimization.^47–51^ Our findings provide further insights on how adherence to a personalized prehab program increases functional reserve, encompassing physical strength, cognitive abilities and immune health,^7–10,52^ allowing patients to increase their resilience and better adapt to surgical stress.

This study has several limitations. All patients underwent a prehab regimen, limiting our ability to compare its effect to no intervention. Each patient therefore served as their own control. The heterogeneity of surgical procedures may have introduced variability in postoperative outcomes. Finally, while mass cytometry allows for simultaneous detection of up to 50 parameters at a single-cell level, the technology necessitates preselection of cell-surface and intracellular features. Future studies, including untargeted transcriptomic, proteomic, or metabolomic approaches, are required to provide a more comprehensive profile of patients’ inflammatory, metabolic, and hormonal responses to prehab. Despite these limitations, our study finds strong evidence of immune and clinical changes after personalized prehab, warranting subsequent, more powered trials to validate our findings.

In conclusion, our study demonstrates a significant effect of a personalized prehab regimen on physical, cognitive, and immune function. Importantly, our study identifies key features echoing previous work looking at the prediction of surgical site infection and postoperative neurocognitive decline after surgery^31–33^ that are positively modulated with prehab. Identification of cell-specific immune features reliably modulated in the personalized prehab group provides a promising set of immune biomarkers that may form the foundation for immune-based diagnostics to identify high-risk patients most likely to benefit from personalized prehab. Moreover, these biomarkers may serve as surrogate endpoints to evaluate preoperative readiness and their risk for postoperative complications throughout their preoperative journey. Tailoring prehab programs to individual patients based on objective immunological endpoints will be crucial for the development of data-driven and effective prehab interventions for high-risk surgical patients.

## Data Availability

All data produced in the present study are available upon reasonable request to the authors

## ACKNOWLEDGEMENTS

J.H., B.G., and F.V. are listed as inventors on a patent application (PCT/US2023/074903). J.H., B.G., D.G., and F.V. are advisory board members, BC is employed, and E.G. is a consultant at SurgeCare SAS. The remaining authors declare no competing interests.

## REFERENCES

1. Tevis SE, Kennedy GD. Postoperative complications and implications on patient-centered outcomes. J Surg Res. 2013;181(1):106–113. doi:10.1016/j.jss.2013.01.032

2. GlobalSurg Collaborative and National Institute for Health Research Global Health Research Unit on Global Surgery. Global variation in postoperative mortality and complications after cancer surgery: a multicentre, prospective cohort study in 82 countries. Lancet. 2021;397(10272):387–397. doi:10.1016/S0140-6736(21)00001-5

3. Pearse RM, Moreno RP, Bauer P, et al. Mortality after surgery in Europe: a 7 day cohort study. Lancet. 2012;380(9847):1059–1065. doi:10.1016/S0140-6736(12)61148-9

4. O’Brien WJ, Gupta K, Itani KMF. Association of Postoperative Infection With Risk of Long-term Infection and Mortality. JAMA Surgery. 2020;155(1):61–68. doi:10.1001/jamasurg.2019.4539

5. Patel AS, Bergman A, Moore BW, Haglund U. The economic burden of complications occurring in major surgical procedures: a systematic review. Appl Health Econ Health Policy. 2013;11(6):577–592. doi:10.1007/s40258-013-0060-y

6. Wilmore DW, Kehlet H. Management of patients in fast track surgery. BMJ. 2001;322(7284):473–476.

7. Carli F, Scheede-Bergdahl C. Prehabilitation to enhance perioperative care. Anesthesiol Clin. 2015;33(1):17–33. doi:10.1016/j.anclin.2014.11.002

8. Carli F, Zavorsky GS. Optimizing functional exercise capacity in the elderly surgical population. Current Opinion in Clinical Nutrition & Metabolic Care. 2005;8(1):23.

9. Humeidan ML, Reyes JPC, Mavarez-Martinez A, et al. Edect of Cognitive Prehabilitation on the Incidence of Postoperative Delirium Among Older Adults Undergoing Major Noncardiac Surgery: The Neurobics Randomized Clinical Trial. JAMA Surg. 2021;156(2):148–156. doi:10.1001/jamasurg.2020.4371

10. Cambriel A, Choisy B, Hedou J, et al. Impact of preoperative uni- or multimodal prehabilitation on postoperative morbidity: meta-analysis. BJS Open. 2023;7(6):zrad129. doi:10.1093/bjsopen/zrad129

11. Carli F, Bousquet-Dion G, Awasthi R, et al. Edect of Multimodal Prehabilitation vs Postoperative Rehabilitation on 30-Day Postoperative Complications for Frail Patients Undergoing Resection of Colorectal Cancer: A Randomized Clinical Trial. JAMA Surgery. 2020;155(3):233–242. doi:10.1001/jamasurg.2019.5474

12. McIsaac DI, Gill M, Boland L, et al. Prehabilitation in adult patients undergoing surgery: an umbrella review of systematic reviews. Br J Anaesth. 2022;128(2):244–257. doi:10.1016/j.bja.2021.11.014

13. Hughes MJ, Hackney RJ, Lamb PJ, Wigmore SJ, Christopher Deans DA, Skipworth RJE. Prehabilitation Before Major Abdominal Surgery: A Systematic Review and Meta-analysis. World J Surg. 2019;43(7):1661–1668. doi:10.1007/s00268-019-04950-y

14. Marmelo F, Rocha V, Moreira-Gonçalves D. The impact of prehabilitation on post-surgical complications in patients undergoing non-urgent cardiovascular surgical intervention: Systematic review and meta-analysis. Eur J Prev Cardiol. 2018;25(4):404–417. doi:10.1177/2047487317752373

15. Moran J, Guinan E, McCormick P, et al. The ability of prehabilitation to influence postoperative outcome after intra-abdominal operation: A systematic review and meta-analysis. Surgery. 2016;160(5):1189–1201. doi:10.1016/j.surg.2016.05.014

16. Daniels SL, Lee MJ, George J, et al. Prehabilitation in elective abdominal cancer surgery in older patients: systematic review and meta-analysis. BJS Open. 2020;4(6):1022–1041. doi:10.1002/bjs5.50347

17. Waterland JL, McCourt O, Edbrooke L, et al. Edicacy of Prehabilitation Including Exercise on Postoperative Outcomes Following Abdominal Cancer Surgery: A Systematic Review and Meta-Analysis. Front Surg. 2021;8:628848. doi:10.3389/fsurg.2021.628848

18. Cohen ME, Liu Y, Ko CY, Hall BL. An Examination of American College of Surgeons NSQIP Surgical Risk Calculator Accuracy. J Am Coll Surg. 2017;224(5):787–795.e1. doi:10.1016/j.jamcollsurg.2016.12.057

19. Bilimoria KY, Liu Y, Paruch JL, et al. Development and evaluation of the universal ACS NSQIP surgical risk calculator: a decision aid and informed consent tool for patients and surgeons. J Am Coll Surg. 2013;217(5):833–842.e1-3. doi:10.1016/j.jamcollsurg.2013.07.385

20. Liu Y, Cohen ME, Hall BL, Ko CY, Bilimoria KY. Evaluation and Enhancement of Calibration in the American College of Surgeons NSQIP Surgical Risk Calculator. J Am Coll Surg. 2016;223(2):231–239. doi:10.1016/j.jamcollsurg.2016.03.040

21. Vos EL, Russo AE, Hohmann A, et al. Performance of the American College of Surgeons NSQIP Surgical Risk Calculator for Total Gastrectomy. J Am Coll Surg. 2020;231(6):650–656. doi:10.1016/j.jamcollsurg.2020.09.023

22. Angele MK, Faist E. Clinical review: Immunodepression in the surgical patient and increased susceptibility to infection. Crit Care. 2002;6(4):298–305.

23. Ni Choileain N, Redmond HP. Cell response to surgery. Arch Surg. 2006;141(11):1132–1140. doi:10.1001/archsurg.141.11.1132

24. Stoecklein VM, Osuka A, Lederer JA. Trauma equals danger--damage control by the immune system. J Leukoc Biol. 2012;92(3):539–551. doi:10.1189/jlb.0212072

25. Liangos O, Domhan S, Schwager C, et al. Whole blood transcriptomics in cardiac surgery identifies a gene regulatory network connecting ischemia reperfusion with systemic inflammation. PLoS One. 2010;5(10):e13658. doi:10.1371/journal.pone.0013658

26. Gaudillière B, Fragiadakis GK, Bruggner RV, et al. Clinical recovery from surgery correlates with single-cell immune signatures. Sci Transl Med. 2014;6(255):255ra131. doi:10.1126/scitranslmed.3009701

27. Ganio EA, Stanley N, Lindberg-Larsen V, et al. Preferential inhibition of adaptive immune system dynamics by glucocorticoids in patients after acute surgical trauma. Nat Commun. 2020;11(1):3737. doi:10.1038/s41467-020-17565-y

28. Huber-Lang M, Lambris JD, Ward PA. Innate immune responses to trauma. Nat Immunol. 2018;19(4):327–341. doi:10.1038/s41590-018-0064-8

29. Wynn TA, Vannella KM. Macrophages in Tissue Repair, Regeneration, and Fibrosis. Immunity. 2016;44(3):450–462. doi:10.1016/j.immuni.2016.02.015

30. Sugimoto MA, Vago JP, Perretti M, Teixeira MM. Mediators of the Resolution of the Inflammatory Response. Trends Immunol. 2019;40(3):212–227. doi:10.1016/j.it.2019.01.007

31. Hédou J, Marić I, Bellan G, et al. Discovery of sparse, reliable omic biomarkers with Stabl. Nat Biotechnol. Published online January 2, 2024:1–13. doi:10.1038/s41587-023-02033-x

32. Verdonk F, Cambriel A, Hedou J, et al. An immune signature of postoperative cognitive decline: A prospective cohort study. Int J Surg. Published online October 16, 2024. doi:10.1097/JS9.0000000000002118

33. Rumer KK, Hedou J, Tsai A, et al. Integrated Single-cell and Plasma Proteomic Modeling to Predict Surgical Site Complications: A Prospective Cohort Study. Ann Surg. 2022;275(3):582–590. doi:10.1097/SLA.0000000000005348

34. Citations – REDCap. Accessed October 9, 2024. https://projectredcap.org/resources/citations/

35. Clavien PA, Barkun J, de Oliveira ML, et al. The Clavien-Dindo Classification of Surgical Complications: Five-Year Experience. Annals of Surgery. 2009;250(2):187–196. doi:10.1097/SLA.0b013e3181b13ca2

36. Barberan-Garcia A, Ubré M, Roca J, et al. Personalised Prehabilitation in High-risk Patients Undergoing Elective Major Abdominal Surgery: A Randomized Blinded Controlled Trial. Ann Surg. 2018;267(1):50–56. doi:10.1097/SLA.0000000000002293

37. Blumenau Pedersen M, Saxton J, Birch S, Rasmussen Villumsen B, Bjerggaard Jensen J. The use of digital technologies to support home-based prehabilitation prior to major surgery: A systematic review. The Surgeon. 2023;21(6):e305–e315. doi:10.1016/j.surge.2023.05.006

38. Ormel H l., van der Schoot G g. f., Sluiter W j., Jalving M, Gietema J a., Walenkamp A m. e. Predictors of adherence to exercise interventions during and after cancer treatment: A systematic review. Psycho-Oncology. 2018;27(3):713–724. doi:10.1002/pon.4612

39. Teixeira PJ, Carraça EV, Markland D, Silva MN, Ryan RM. Exercise, physical activity, and self-determination theory: A systematic review. International Journal of Behavioral Nutrition and Physical Activity. 2012;9(1):78. doi:10.1186/1479-5868-9-78

40. Carli F, Baldini G, Feldman LS. Redesigning the Preoperative Clinic: From Risk Stratification to Risk Modification. JAMA Surg. 2021;156(2):191–192. doi:10.1001/jamasurg.2020.5550

41. Aiello A, Ligotti ME, Garnica M, et al. How Can We Improve Vaccination Response in Old People? Part I: Targeting Immunosenescence of Innate Immunity Cells. Int J Mol Sci. 2022;23(17):9880. doi:10.3390/ijms23179880

42. Fulop T, McElhaney J, Pawelec G, et al. Frailty, Inflammation and Immunosenescence. Published online July 20, 2015. doi:10.1159/000381134

43. Qian F, Wang X, Zhang L, et al. Age-associated elevation in TLR5 leads to increased inflammatory responses in the elderly. Aging Cell. 2012;11(1):104–110. doi:10.1111/j.1474-9726.2011.00759.x

44. Tavenier J, Rasmussen LJH, Houlind MB, et al. Alterations of monocyte NF-κB p65/RelA signaling in a cohort of older medical patients, age-matched controls, and healthy young adults. Immunity & Ageing. 2020;17(1):25. doi:10.1186/s12979-020-00197-7

45. Lin HS, Watts JN, Peel NM, Hubbard RE. Frailty and post-operative outcomes in older surgical patients: a systematic review. BMC Geriatr. 2016;16(1):157. doi:10.1186/s12877-016-0329-8

46. Shinall MC Jr, Arya S, Youk A, et al. Association of Preoperative Patient Frailty and Operative Stress With Postoperative Mortality. JAMA Surgery. 2020;155(1):e194620. doi:10.1001/jamasurg.2019.4620

47. Shao T, Verma HK, Pande B, et al. Physical Activity and Nutritional Influence on Immune Function: An Important Strategy to Improve Immunity and Health Status. Front Physiol. 2021;12. doi:10.3389/fphys.2021.751374

48. Gillis C, Wischmeyer PE. Pre-operative nutrition and the elective surgical patient: why, how and what? Anaesthesia. 2019;74(S1):27–35. doi:10.1111/anae.14506

49. Childs CE, Calder PC, Miles EA. Diet and Immune Function. Nutrients. 2019;11(8):1933. doi:10.3390/nu11081933

50. Dantzer R. Neuroimmune Interactions: From the Brain to the Immune System and Vice Versa. Physiol Rev. 2018;98(1):477–504. doi:10.1152/physrev.00039.2016

51. Lyu D wei. Immunomodulatory edects of exercise in cancer prevention and adjuvant therapy: a narrative review. Front Physiol. 2024;14. doi:10.3389/fphys.2023.1292580

52. Skořepa P, Ford KL, Alsuwaylihi A, et al. The impact of prehabilitation on outcomes in frail and high-risk patients undergoing major abdominal surgery: A systematic review and meta-analysis. Clin Nutr. 2024;43(3):629–648. doi:10.1016/j.clnu.2024.01.020

53. Fragiadakis GK, Gaudillière B, Ganio EA, et al. Patient-specific Immune States before Surgery Are Strong Correlates of Surgical Recovery. Anesthesiology. 2015;123(6):1241–1255. doi:10.1097/ALN.0000000000000887

