## Supplementary materials for "Immune Modulation by Personalized vs. Standard Prehabilitation Before Major Surgery: A Randomized Controlled Trial"

Amelie Cambriel et al

This PDF files includes

Supplementary methods

Figures S1 to S3

Table S1 to S2

**Supplementary methods:**

*Single cell analysis*

*Blood collection*

Whole blood samples were collected at enrollment, i.e. before prehab, and on the day of surgery before anesthesia. They were collected in sodium heparinized tubes. Then, samples were either left unstimulated to measure endogenous intracellular activities or exposed to either the toll like receptor (TLR) 4 agonist lipopolysaccharide (LPS), or Inter-leukine (IL) 2,4 and 6, or Tumor necrosis factor alpha (TNFα) to activate canonical surgical trauma related signaling responses in vitro. This in vitro *immune stress test* aims to replicate surgical trauma in vitro and provide an in depth analysis of patients' immunome[32,33] (**Figure 1B**). All samples were then fixed with Proteomic Stabilizer in Smart Tubes (Smart α Inc., San Carlos, CA) and immediately stored at -80˚C.

*Barcoding, antibody staining and mass cytometry processing*

Following erythrocyte lysis, samples were barcoded and stained with both surface and intracellular antibodies according to standard protocols[26,27]. The antibody panel comprised 41-marker including 26 antibodies targeting cell-surface markers and 15 intracellular antibodies specific to phosphorylated (p) signaling epitopes (**Table S1**). The selection of immune cell types and intracellular signaling pathways was informed by previous studies on the peripheral immune response to surgery [27,32,34] and preclinical evidence related to immune processes involved in post-operative complications, particularly surgical site infection (SSI) and postoperative neurocognitive decline (POND) [35]. To reduce variability in measurement, samples from each individual patient for a specific stimulation condition were processed simultaneously. The resulting FCS files were normalized and de-barcoded using MatLab-based software, then uploaded to the Cell Engine platform (https://cellengine.com, Primity Bio, Fremont, CA) for manual gating.

*Cell frequency, endogenous intracellular signaling, and intracellular signaling responses*

For each patient, we analyzed 1096 single-cell proteomic features per timepoint. Features included the frequency of 41 key innate and adaptive immune cells defined through manual gating based on pre-established gating strategies [27] (**Fig. S1**), as well as their intracellular signaling activities (e.g., the phosphorylation state of 11 proteins including pSTAT1, pSTAT3, pSTAT5, pSTAT6, pNF-κB, pMAPKAPK2, pP38, prpS6, pERK1/2, pCREB, and total IκBα). Immune cell frequency features were calculated for each immune cell subset from the unstimulated samples. For each cell type, intracellular activities were reported as the median signal intensity (arcsinh transformed value). Changes in signaling in response to receptor-specific ligands were expressed as the arcsinh transformed ratio relative to the endogenous signaling, i.e., the difference in arcsinh transformed signal intensity between the stimulated and unstimulated condition. The proportion of mononuclear cells was reported as a percentage of the gated live mononuclear cells. The proportion of neutrophils was reported as a percentage of the gated live leukocytes.

*Preprocessing and Visualization of the single-cell proteomic data set*

Before conducting statistical analysis, three preprocessing steps were applied to the initial dataset. First, a knowledge-based penalization matrix was applied to the intracellular signaling response features in the mass cytometry data based on mechanistic immunological knowledge, as previously described[33]. Second, features that exhibited no variance were excluded from the dataset. Lastly, features were Z-scored before fitting any model. The single-cell mass cytometry dataset was then visualized using a uniform manifold approximation and projection (UMAP) layout.

*Stabl algorithm*

Stabl is a supervised machine learning framework that allows for sparse, reliable, and predictive feature selection[33]. Stabl is particularly well-suited for this study, given the relatively small number of observations (n) compared to the number of features (p). It yields a sparse solution that builds the final model from a limited number of selected features. The package for Stabl is available online.

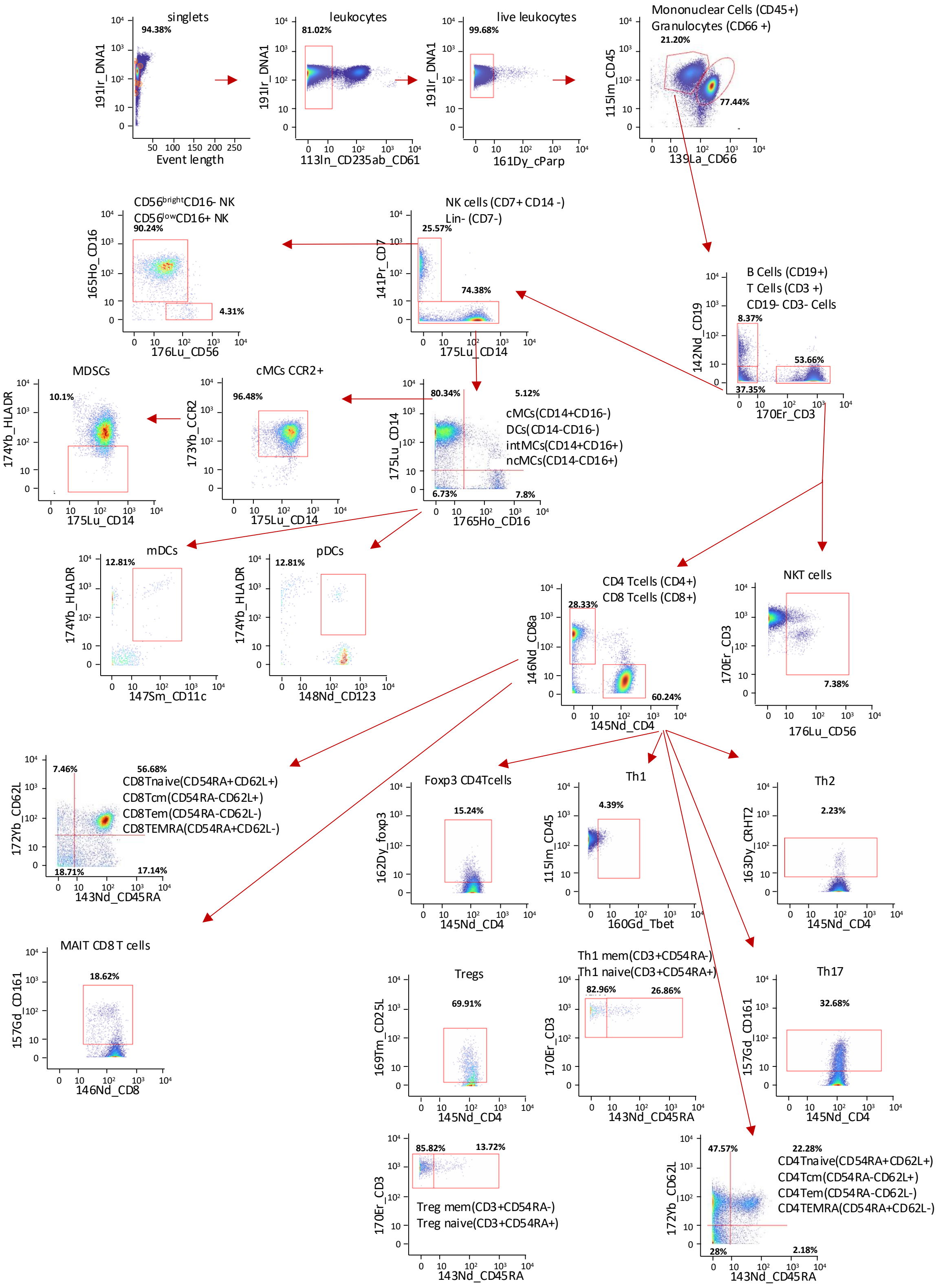


**Figure S1:** Immune cell gating strategy

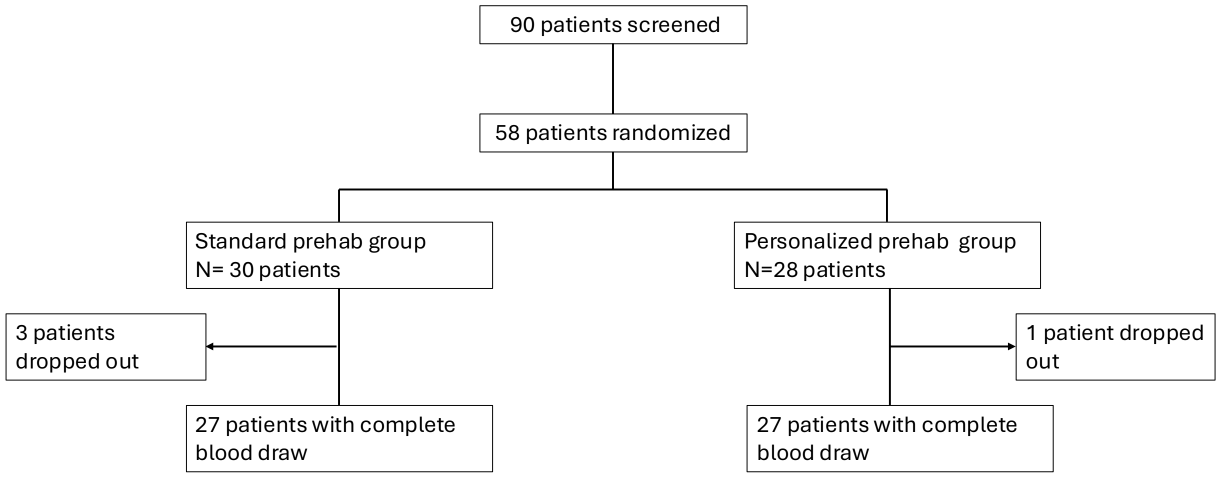


**Figure S2:** Study flow chart according to CONSORT guidelines.

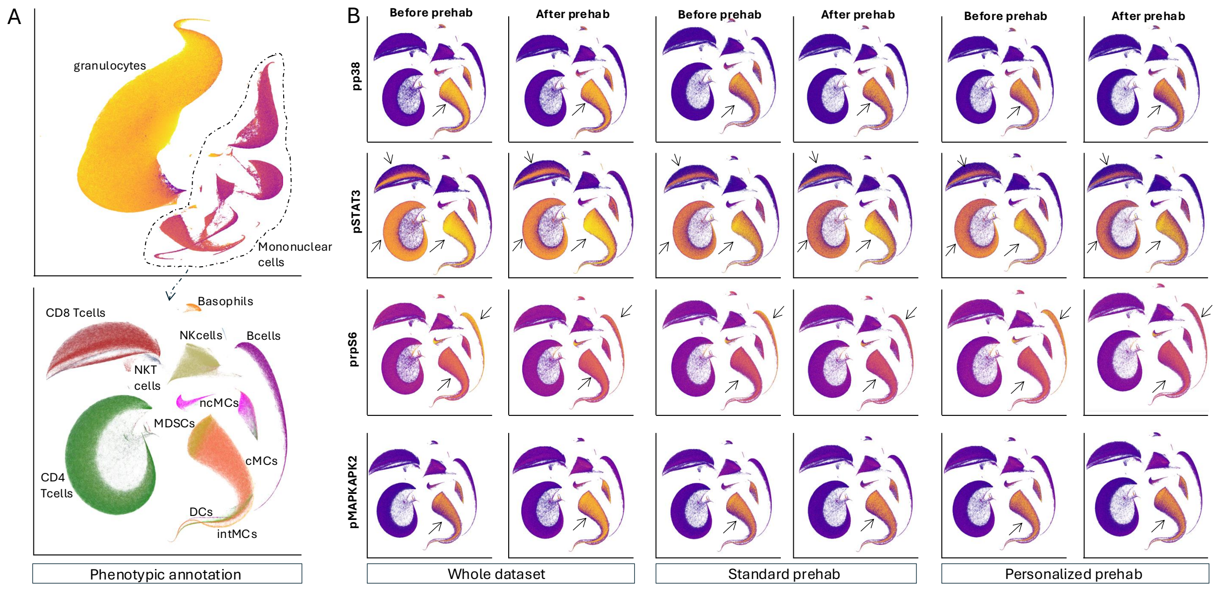


**Figure S3: A.** UMAP representation of the single-cell mass cytometry dataset. Top, all live leukocytes, including neutrophils and mononuclear cells; Bottom: UMAP representation of mononuclear cells only. UMAPS are clustered by cell types and annotated. **B.** UMAPs representing all mononuclear cells colored according intra-cellular signaling before and after prehab; left for the whole dataset, middle for the standard prehab group, right for the personalized prehab group *Abbreviations: cMCs: classical monocytes; CREB: cAMP-response element binding protein; intMCs: intermediate monocytes; mDCs: myeloid dendritic cells; MAPKAPK2: Mitogen-activated protein kinase-activated protein kinase ;, MDSCs: Myeloid Derived Suppressor Cells; NF-κB: Nuclear factor – κB; ncMCs: non classical monocytes; NK cells: natural killer cells; pDCs: plasmacytoid dendritic cells; p: phosphorylation; STAT: Janus Kinase/signal transducers and activators of transcriptions; TNFα: Tumor Necrosis Factor alpha*

.

| Antibody | Manufacturer | Metal | Isotope | Clone | Concentration | Comment |
|---|---|---|---|---|---|---|
| **Barcode1** | Trace Sciences | Pd | 102 | | 15μM | Barcode |
| **Barcode2** | Trace Sciences | Pd | 104 | | 15μM | Barcode |
| **Barcode3** | Trace Sciences | Pd | 105 | | 15μM | Barcode |
| **Barcode4** | Trace Sciences | Pd | 106 | | 15μM | Barcode |
| **Barcode5** | Trace Sciences | Pd | 108 | | 15μM | Barcode |
| **Barcode6** | Trace Sciences | Pd | 110 | | 15μM | Barcode |
| **CD235ab** | Biolegend | In | 113 | HIR2 | 1μg/mL | Phenotype |
| **CD61** | BD | In | 113 | VI-PL2 | 0.5μg/mL | Phenotype |
| **CD45** | Biolegend | In | 115 | HI30 | 1μg/mL | Phenotype |
| **CD66** | BD | La | 139 | CD66a-B1.1 | 0.5μg/mL | Phenotype |
| **CD7** | BD | Pr | 141 | M-T701 | 0.5μg/mL | Phenotype |
| **CD19** | Biolegend | Nd | 142 | HIB19 | 0.5μg/mL | Phenotype |
| **CD45RA** | Biolegend | Nd | 143 | HI100 | 0.5μg/mL | Phenotype |
| **CD11b** | Biolegend | Nd | 144 | ICRF44 | 2μg/mL | Phenotype |
| **CD4** | Biolegend | Nd | 145 | RPA-T4 | 2μg/mL | Phenotype |
| **CD8a** | Biolegend | Nd | 146 | RPA-T8 | 1μg/mL | Phenotype |
| **CD11c** | Biolegend | Sm | 147 | Bu15 | 1μg/mL | Phenotype |
| **CD123** | Biolegend | Nd | 148 | 6H6 | 1μg/mL | Phenotype |
| **pCREB** | Cell Signaling Technology | Sm | 149 | 87G3 | 2μg/mL | Function |
| **pSTAT5** | Cell Signaling Technology | Nd | 150 | C11C5 | 4μg/mL | Function |
| **pP38** | BD | Eu | 151 | 36/p38 | 2μg/mL | Function |
| **TCRgd** | BD | Sm | 152 | B1 | 4μg/mL | Phenotype |
| **pSTAT1** | BD | Eu | 153 | 14/P-STAT1 | 1μg/mL | Function |
| **pSTAT3** | Cell Signaling Technology | Sm | 154 | M9C6 | 2μg/mL | Function |
| **pS6** | Cell Signaling Technology | Gd | 155 | D57.2.2E | 2μg/mL | Function |
| **FceRI** | Biolegend | Gd | 156 | AER-37 (CRA-1) | 0.5 μg/mL | Phenotype |
| **CD33** | Biolegend | Gd | 158 | WM53 | 2μg/mL | Phenotype |
| **pMAPKAPK2** | Cell Signaling Technology | Tb | 159 | 27B7 | 1μg/mL | Function |
| **Tbet** | Thermo Fisher | Gd | 160 | 4B10 | 8μg/mL | Phenotype |
| **cPARP** | BD | Dy | 161 | F21-852 | 1μg/mL | Phenotype |
| **FoxP3** | Thermo Fisher | Dy | 162 | PCH101 | 8μg/mL | Phenotype |
| **IκB** | Cell Signaling Technology | Dy | 164 | L35A5 | 8μg/mL | Function |
| **CD16** | Biolegend | Ho | 165 | 3G8 | 1μg/mL | Phenotype |
| **pNF-κB** | BD | Er | 166 | K10-895.12.50 | 2μg/mL | Function |
| **pERK1-2** | Cell Signaling Technology | Er | 167 | D13.14.4E | 4μg/mL | Function |
| **pSTAT6** | Biolegend | Er | 168 | A15137E | 1μg/mL | Function |
| **CD25** | Biolegend | Tm | 169 | M-A251 | 2μg/mL | Phenotype |
| **CD3** | Biolegend | Er | 170 | UCHT1 | 1μg/mL | Phenotype |
| **CXCR4** | BD | Yb | 171 | M-T271 | 2μg/mL | Phenotype |
| **CD62L** | Biolegend | Yb | 172 | W6D3 | 0.5μg/mL | Phenotype |
| **CCR2** | Biolegend | Yb | 173 | K036C2 | 2μg/mL | Phenotype |
| **HLA-DR** | Fluidigm | Yb | 174 | L243 | 2μg/mL | Phenotype |
| **CD14** | Fluidigm | Lu | 175 | M5E2 | 2μg/mL | Phenotype |
| **CD56** | BD | Lu | 176 | NCAM16.2 | 1μg/mL | Phenotype |
| **DNA1** | Fluidigm | Ir | 191 | | 50μM | DNA |
| **DNA2** | Fluidigm | Ir | 193 | | 50μM | DNA |

**Table S1:** Mass Cytometry antibody panel

| | Standard prehab N=27 | Personalized prehab N=27 | p |
|---|---|---|---|
| **Patients with Clavien-Dindo complication grade >1** | 11 (40.7) | 4 (15.8) | **0.037**[1] |
| **Complication grade at POD30 according to Clavien-Dindo classification** | | | |
| Grade 0 | 0 (0) | 2 (7.1) | |
| Grade 1 | 18 (64.3) | 21 (75.0) | |
| Grade 2 | 8(28.6) | 3(10.7) | |
| Grade 3a | 2(7.1) | 0 (0) | |
| Grade 3b | 1(3.5) | 1 (3.5) | |
| **Hospital length of stay, days** | 2.6[1.5,6.03] | 2.12[0.66,4.02] | 0.48[2] |

[1]Mann-Whitney Utest

[2]Wilcoxon rank sum test

**Table S2:** Postoperative outcomes
